# Immune responses after twofold SARS-CoV-2 immunisation in elderly residents and Health Care Workers in nursing homes and homes with assisted living support - Proposal for a correlate of protection

**DOI:** 10.1101/2022.02.09.22270747

**Authors:** Julia Schiffner, Nora Eisemann, Hannah Baltus, Sina Jensen, Katharina Wunderlich, Stefan Schüßeler, Charlotte Eicker, Bianca Teegen, Doreen Boniakowsky, Werner Solbach, Alexander Mischnik

**Author notes:** **Corresponding author:** Alexander Mischnik, M.D., Health Protection Authority, Sophienstr. 2-8, D-23560 Luebeck, Germany. shared first authorship (Julia Schiffner, Nora Eisemann). shared last authorship (Werner Solbach, Alexander Mischnik).

## Abstract

In the present study, we were interested in the decline over time of anti SARS-CoV-2 antibodies and SARS-CoV-2 specific T-cell responses after two doses of mRNA vaccines in total and by age group and comorbidity. The second goal was to suggest an immunological correlate for protection on an individual basis and to describe the probability of protection over time after second vaccination.

We analysed blood samples from 228 residents (median age 83.8 years) and from 273 Health Care Workers (HCW; median age 49.7 years) of five nursing homes and one home for the elderly with assisted living support. Participants had received two vaccinations. The blood samples were analysed for SARS-CoV-2 specific antibody and T-cell responses. We compared outcomes in the HCW and residents in the respective institutions. No breakthrough infections occurred during the study period. The initial immune responses in the younger participants were about 30 % higher than in the older ones. Over time, all parameters dropped continuously in all groups within the maximum observation period of 232 days. Comorbidities such as coronary heart disease or diabetes mellitus reduced the initial immune responses, regardless of age. In contrast to an almost linear decline in antibody levels, we observed that the interferon-gamma response remained at a constant level between about day 120 and 180, only to decline further thereafter.

Based on our data, we propose on an individual level a correlate of protection: Persons who have a neutralizing capacity of 75 % (which would correspond to approx. 200 BAU/ml) and an interferon-gamma response above 200 mIU/ml should be considered to be protected resp. sufficiently immunized.

## Introduction

Multiple vaccines have been developed that offer protection against COVID-19 by generating immune responses against the spike antigen of SARS-CoV-2. On 27 December 2020, the national vaccination program started in Germany with the Pfizer–BioNTech BNT162b2 mRNA vaccine (B/P Comirnaty) followed by the approval of Spikevax mRNA (mRNA; Moderna) at 6^th^ January 2021 and ChAdOx1 nCoV-19 vector vaccine (Vaxzevria; Astra Zeneca AZ) which was first recommended on 29^th^ January 2021 (1). Initially, vaccines were administered to priority groups, including residents of old people’s and nursing homes, persons aged ≥ 80 years, personnel with a particularly high risk of exposure in medical facilities (e.g. in emergency rooms, in the medical care of COVID-19 patients), personnel in medical facilities with close contact to vulnerable groups (e.g. in oncology or transplant medicine), nursing staff in outpatient and inpatient care for the elderly, other workers in homes for the elderly and nursing homes with contact to residents. Later, vaccines were recommended population-wide for anybody ≥ 12 years.

It has been reported by us (2), (3) and others (4-6) that the cellular and humoral immune response wanes over time after infection and after vaccination. In the present study, we were primarily interested how anti SARS-CoV-2 antibodies and SARS-CoV-2 specific T-cell responses declined over time after two doses of mRNA vaccines, especially by age-group and comorbidity status. The second goal was to set up a proposal for an immunological correlate of protection on an individual basis which appears sensible on the basis of the the given data.

In the present study we analysed blood samples from 228 residents (median age 83.8 years) and from 273 Health Care Workers (HCW; median age 49.7 years) of five nursing homes and one home for the elderly with assisted living support. Participants had received two vaccinations (mostly Pfizer/BioNTech BNT162b2 mRNA vaccine (B/P Comirnaty). The blood samples were analysed for SARS-CoV-2 specific antibody and T-cell responses. We compared outcomes in younger persons (< 65 years, mostly health care workers (HCW)) and in older persons (65 years or older, mostly residents in the respective institutions). The study period was between 31^st^ August and 9^th^ September 2021. At this time the Delta variant prevailed (98 %), the first Omikron variant appeared appeared in the region on 14^th^ December 2021.

We suggest a rationale to support the decision if an individual is sufficiently protected against severe courses of disease and describe how the protection status changes over time. The present recommendations for vaccination do not rely on individual reactivity of the immune system but solely on protection data in large cohorts. Missing is a clinical parameter to support practioners in deciding if an individual is protected or not. The need of a booster vaccination is extensively documented in various studies (7), but the currently recommended timepoint – from three months after the second vaccination - might not be appropriate for the individual person.

## Material and methods

### Study population

Persons either living or working in six old people’s homes in Northern Germany were recruited. Five facilities are stationary retirement homes and one facilitiy is a so called assisted living home. In total, 1,228 persons were invited by e-mail or personal contact to participate. Inclusion criteria were being vaccinated twice against SARS-CoV-2, an elapsed period of at least 14 days since the second vaccination (as the vaccination effect is first built up during this time) and written informed consent. Exclusion criteria were a third vaccination, unknown date of second vaccination, unsuccessful blood drawings, no laboratory result of the blood sample or a positive test for anti-SARS-CoV-2 nucleocapsid protein (NCP) antibodies. Such a positive test may indicate an undetected infection, which would bias the results. **Figure 1** shows the flow-chart of recruitment. Of the final 501 study participants, 273 were health care workers (HCW in the retirement homes) and 228 were seniors living in the facilities.

**Figure 1:**
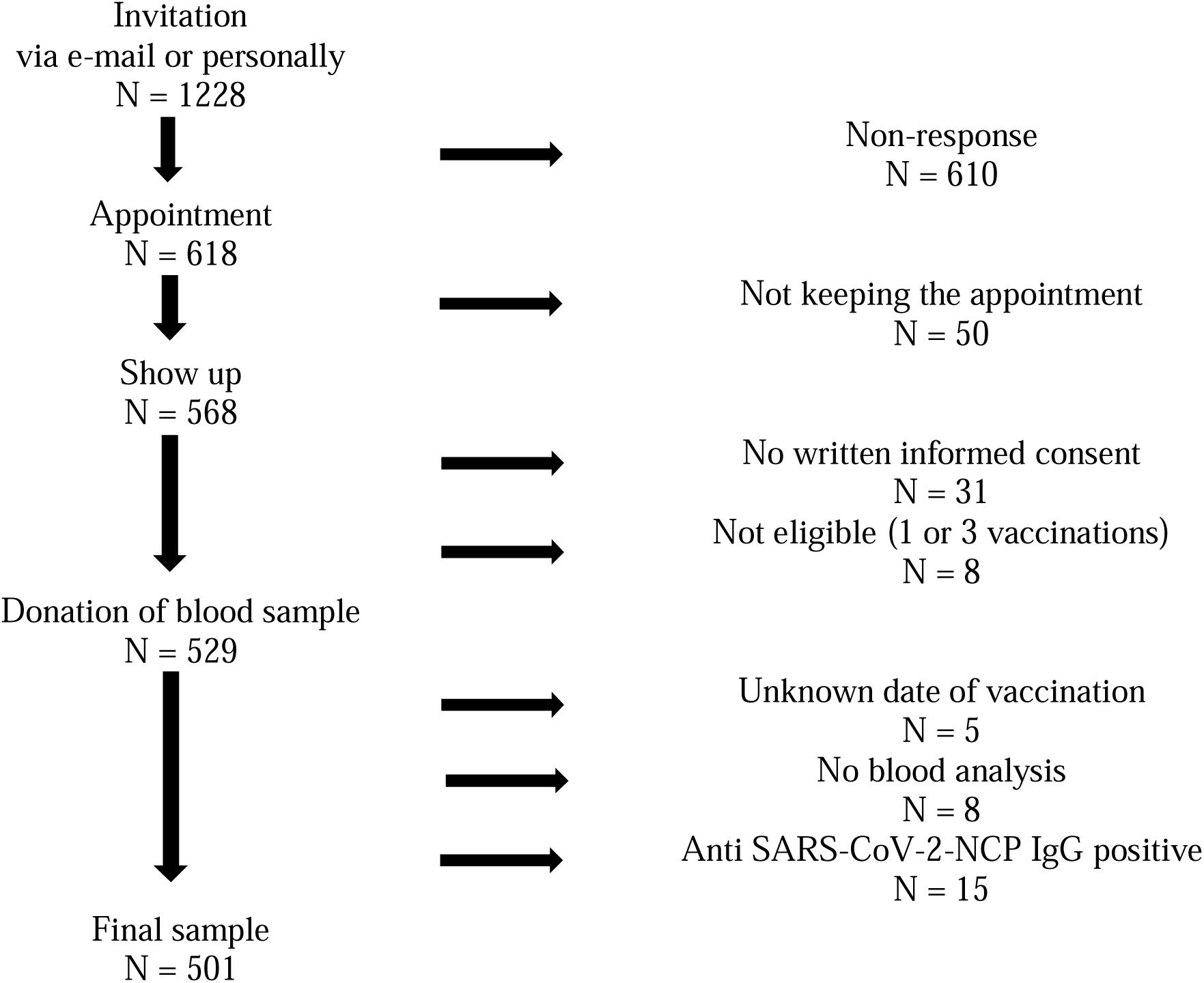
Flow chart of the ISCOV-VAC study recruitment.

At the study visit, blood samples were taken from the participants and transferred directly to the laboratory within four hours. In addition, the participants filled in a questionnaire about personal data (e.g. age, sex, body height and body weight) and comorbidities (such as diabetes, autoimmune diseases, cardio-vasculary disease).

### Laboratory methods

The blood samples were analysed for four main outcomes: anti-SARS-CoV-2 S1-Protein IgG antibodies, antibody neutralization capacity, SARS-CoV-2 S1 reactive T cells (i. e. interferon-gamma release assay, IGRA) and anti-SARS-CoV-2 nucleocapsid protein antibodies.

#### Detection of anti-SARS-CoV-2 S1-Protein IgG antibodies

Serum IgG antibodies against the viral (strain Wuhan-1) S1 domain of the spike protein including the receptor binding domain (RBD) were detected by using the “Anti-SARS-CoV-2 QuantiVac ELISA” detection kit (EUROIMMUN; Lübeck, Germany, product no. EI 2606-9601-10 G) according to the instructions of the manufacturer. The measured “relative units/ml” were calibrated with the “First WHO standard of anti-SARS-CoV-2 immunoglobulin” (NIBSC code 20/136) and converted into Binding Antibody Units (BAU)/ml by multiplication with the factor 3.2. Interpretation is as follows: < 25.6 BAU/ml = negative; ≥ 25.6 BAU/ml = positive.

#### Detection of antibody neutralization capacity

Antibody binding of SARS-CoV-2-S1/RBD neutralizing antibodies was detected by applying the “Anti-SARS-CoV-2 NeutraLISA” detection kit (EUROIMMUN; Lübeck, Germany, product no. EI 2606-9601-4) according to the instructions of the manufacturer. This is a surrogate neutralization test which has 98.6 % concordance when compared to plaque-reduction (PRNT_50_) testing. Specificity is 99.7 % and sensitivity is 95.9 %. Values are interpreted as follows: < 20 % = negative; 20 %-< 35% = borderline; ≥ 35 % = positive.

#### Determination of SARS-CoV-2 S1 reactive T cells (Interferon-gamma release assay, IGRA)

T cells in peripheral blood reacting to SARS-CoV-2 S1 protein were detected by using the “Quant-T-Cell ELISA” (EUROIMMUN; Lübeck, Germany product no. EQ 6841-9601 and ET 2606-3003). In brief, heparinized blood cells were cultured with S1 antigen for 24 hours. Subsequently, Interferon-gamma release was determined in the culture supernatant by ELISA.Values are expressed in mIU/ml. Interpretation is as follows: < 100 mIU/ml = negative, 100 – 200 mIU/ml = borderline; ≥ 200 mIU/ml = positive.

#### Detection of Anti-SARS-CoV-2 nucleocapsid protein (NCP) antibodies

To discriminate between vaccine-induced antibody response and convalescent SARS-CoV-2 infection, serum IgG antibodies against the nucleocapsid protein were detected by using the “Anti-SARS-CoV-2 NCP ELISA” detection kit (EUROIMMUN; Lübeck, Germany product no. EI 2606-9601-2 G). This is a semiquantitative test. Values are given in ratios. Ratios are calculated from the extinction of the sample and that of a standardized calibrator. Interpretation is as follows: Ratio < 0.8 = negative; ≥ 0.8 to < 1.1 = borderline; ≥ 1.1 = positive.

We found 15 positive results. These individuals were excluded from the study. The validity and reliability test characteristics have been described recently (8).

### Statistical methods

Anti-SARS-CoV-2 S1-Protein IgG antibody values above 384 BAU/ml were reported as ‘above 384’ by the laboratory and conservatively set to 385 for further analysis, as was the case with values reported as ‘below 3.2’, which were set to 1.6. Similarly, SARS-CoV-2 S1 reactive T cell values above 2500 mIU/ml were set to 250 mIU/ml. Pairwise correlation coefficients were calculated for SARS-CoV-2 S1 reactive T cells, anti-SARS-CoV-2 S1-Protein IgG antibodies and antibody neutralization capacity. As the Spearman correlation between anti-SARS-CoV-2 S1-Protein IgG antibodies and antibody neutralization capacity was very high, the anti-SARS-CoV-2 S1-Protein IgG antibody data appeared dispensable and we continued only with SARS-CoV-2 S1 reactive T cells and antibody neutralization capacity. The continuous values of interferon-gamma and of neutralizing capacity were each plotted against the number of days since second vaccination. Local polynomial regression models (with linear polynomials) were fitted to describe the change in the outcomes over time, together with the corresponding 95 % prediction intervals. Subgroup analyses by age (age below 65 years versus 65+), sex (female versus male) and comorbidity (no versus any comorbidity) were done. In search for a correlate of protection, we defined the following values as “protective”: antibody neutralization capacity values above 75 % and positive SARS-CoV-2 S1 reactive T cell levels > 200 mIU/ml (rationale for these choices see Discussion section). A log-linear regression model predicted that an antibody neutralization capacity value of 75 % corresponds to an IgG antibody value of about 200 BAU/ml. The estimated proportion of protected individuals together with a 95 % prediction interval is plotted against the number of days since second vaccination. Again, local polynomial regression models (with linear polynomials) were used. All analyses were conducted with R 4.1.1 (9).

### Ethics

The study was conducted in accordance with the Declaration of Helsinki and Good Clinical Practice (10) and approved by the ethics committee of the University of Luebeck (21-353).

## Results

### Characteristics of the study participants

A description of the characteristics of the study population is given in **Table 1**. The age of the participants ranged between 19 and 100 years, with a median age of 83.8 years in residents and 49.7 years in HCW. The majority of residents and HCW was female (67 % and 75 %, respectively). The Body Mass Index (BMI) was in the normal range for 50 % of the residents and 39 % of the HCW, while few participants were underweight (3.9 % and 1.5 % in residents and HCW, respectively) and most were overweight or obese (47 % and 59 % in residents and HCW, respectively). Comorbidities were very common among residents (96 %), but also among HCW (54 %). The most frequent comorbidities were coronary heart disease in residents (79 %) and HCW (25 %) and neurological disease in residents (48 %). Diseases of the immune system (not specified) were mentioned in about 20 % of both residents and HCW.

**Table 1:** Description of the study population.

|  | **Residents** | **Health care workers** |
| --- | --- | --- |
| Total, N (%) | 228 (100 %) | 273 (100 %) |
| Age in years, median (IQR) | 83.8 (79.2 to 88.2)<br>Min: 41.3, max: 100.5 | 49.7 (39.8 to 55.8)<br>Min: 19.5, max: 65.5 |
| Sex female, N (%) | 157 (67.5 %) | 206 (74.7 %) |
| Body mass index, N (%) |  |  |
| Underweight (<18.5) | 9 (3.9 %) | 4 (1.5 %) |
| Normal (18.5 to 25) | 113 (49.6 %) | 107 (39.2 %) |
| Overweight (>25 to 30) | 62 (27.2 %) | 84 (30.8 %) |
| Adipositas (> 30) | 44 (19.3 %) | 78 (28.6 %) |
| Any Comorbidity, N (%) | 215 (96.0 %) | 142 (54.2 %) |
| Coronary heart disease | 177 (79.4 %) | 60 (24.5 %) |
| Diabetes mellitus | 50 (22.8 %) | 17 (7.1 %) |
| Lung disease | 35 (15.8 %) | 28 (11.3 %) |
| Liver disease | 9 (4.0 %) | 1 (0.4 %) |
| Neurological disease | 106 (47.5 %) | 9 (3.7 %) |
| Kidney disease | 56 (25.6 %) | 4 (1.7 %) |
| Cancer | 47 (21.7 %) | 12 (5.0 %) |
| Immune system | 46 (20.9 %) | 50 (19.9 %) |
| Vaccine |  |  |
| AZ & AZ | 3 (1.3 %) | 11 (4.0 %) |
| Biontech/Pfizer | 1 (0.4 %) | 11 (4.0 %) |
| Moderna | - | 1 (0.4 %) |
| unknown | - | 1 (0.4 %) |
| Biontec/Pfizer & B/P | 224 (98.2 %) | 244 (89.4 %) |
| Moderna & Moderna | - | 2 (0.7 %) |
| Unknown & B/P | - | 2 (0.7 %) |
| Unknown | - | 1 (0.4 %) |
| Time between second vaccination and blood sampling, median (IQR) | 195 (150 to 207)<br>Min: 50, max: 224 | 182 (122 to 195)<br>Min: 22, max: 232 |

\* Missing values are excluded from the calculation of the percentage with disease.

Different combinations of vector (Astra Zeneca) and mRNA (B/P and Moderna) vaccines were possible, but the vast majority received B/P as first and second vaccine (98 % in residents and 89 % in HCW). The first vaccination was given between 24th December 2020 and 29th June 2021 and the second between 19th January 2021 and 10th August 2021. The time between second vaccination and blood sampling ranged between 22 and 232 days. No data was collected for the first three weeks after second vaccination because the natural immune response takes about two weeks. We did not observe any symptomatic break-through infection during the observation period.

During the study period, the Delta variant of SARS-CoV-2 was prevailing with more than 98 % of the isolates detected.

### Immune responses

First, we determined how the anti SARS-CoV-2 IgG antibody levels corresponded to the neutralising capacity. The bivariate scatterplot in **Figure 2** shows a high correlation (Spearman correlation of 0.959, 95 % confidence interval [0.951 to 0.966]). Thus, for further considerations we concentrated on the neutralizing capacity. Pairwise scatterplots and correlation coefficients between all three outcomes are given in **Figure 1 in the supplement**. If we look at the dynamics of the neutralising antibodies, **Figure 3** (lower panel) shows an almost linear decrease over the entire study period. After about 200 days after two doses of the vaccine, the neutralisation capacity had dropped from > 90 % to about 40 %.

**Figure 2:**
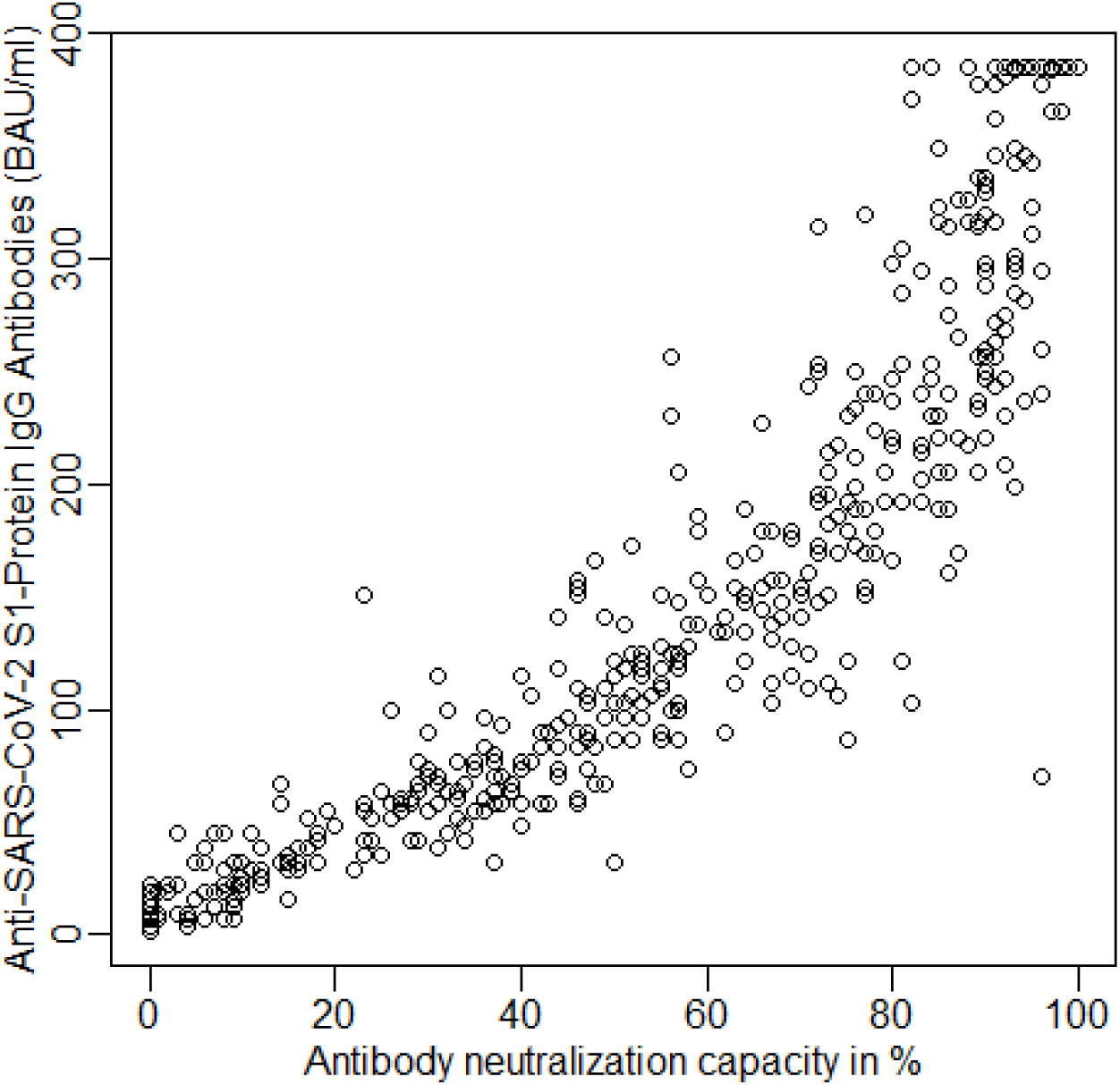
Bivariate scatterplot for Anti-SARS-CoV-2 S1-Protein IgG Antibodies and neutralisation capacity (N=501)

**Figure 3:**
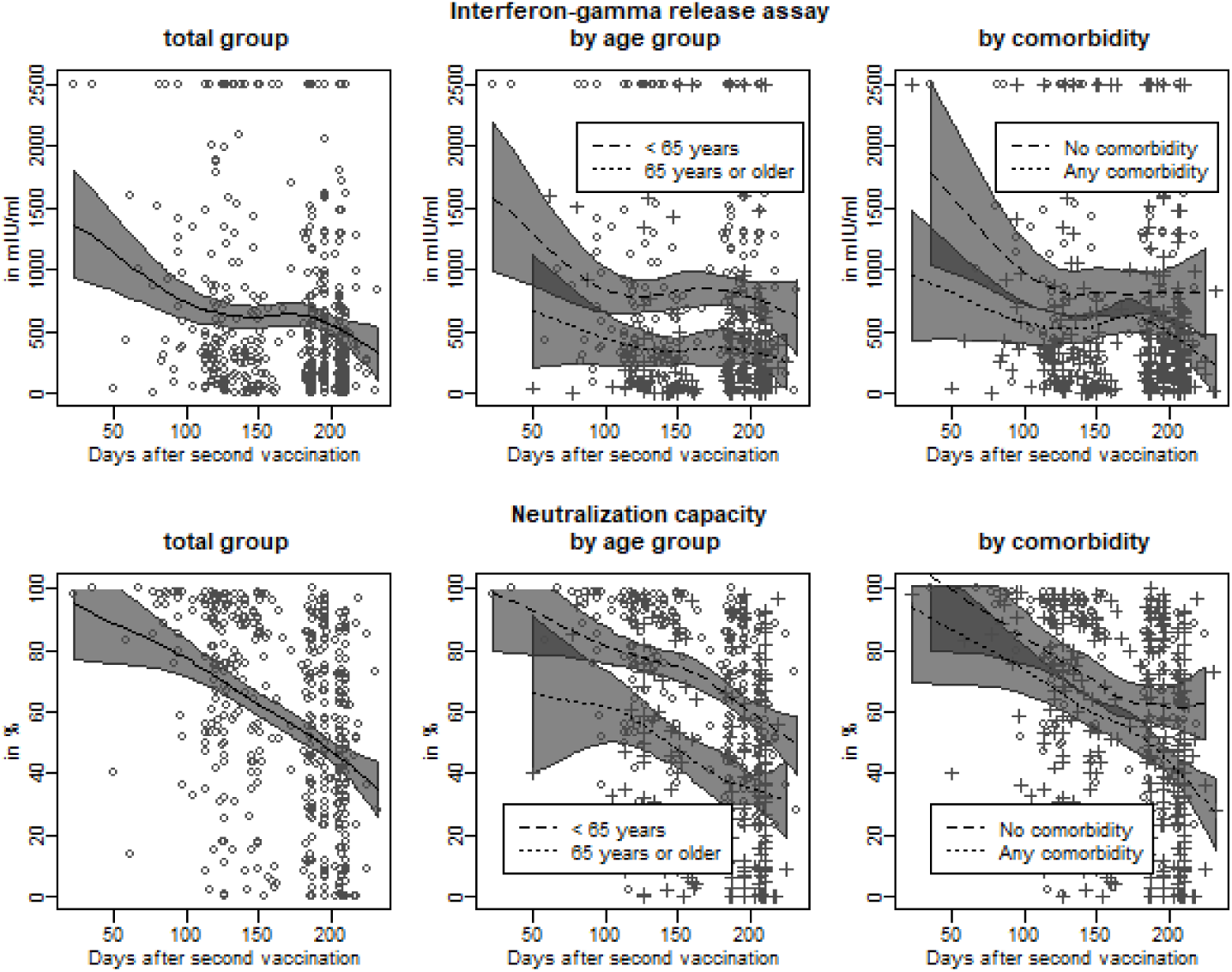
SARS-CoV-2 S1 reactive T cells and neutralisation capacity over time in the total group, by age group and by comorbidity status Legend: Dots – observed values for age up to 65 years or no comorbidity, cross – observed values for age 65+ or any comorbidity, grey area – 95 % prediction band

For the subgroup analysis, we formed two age groups: Persons under 65 years of age, which included mostly the HCWs, and those over 65 years of age, i.e. the residents of the old people’s homes. As expected, the antibody response shortly after the second vaccination was only almost half as high in the elderly group as compared to the younger group (lower panel, middle). However, the relative decline over time was comparable. It is therefore not surprising that the duration of the protective effect of vaccination is limited in the elderly. We were then interested to see how the co-morbidities reported by the subjects were reflected in the antibody response. The predicted neutralisation capacity is slightly lower for persons with existing co-morbidities at any time. The kinetics of the antibody decrease were comparable (lower panel, right). The low number of study participants did not allow to draw any meaningful conclusions on the effect of any single co-morbidity (data not shown). Furthers subgroup analyses did not show relevant differences by sex or by BMI category. Subgroups were too small (and in some cases observations were too differently distributed over time) to make reliable comparisons by care level, vaccine or individual diseases.

When looking at the T-cell response with regard to spike-protein specific interferon-gamma secretion (interferon-gamma release assay, IGRA), it is noticeable that the values also drop linearly between about 50 and about 120 days after the second vaccination, but then remain on a plateau (approx. 700 mIU/ml) until about 180 days, after which they drop further (upper panel, left). Here, too, there is a clear difference between the older (65+) and younger participants (under 65 years). In the elderly, the T-cell reactivity was only about 50 % in comparison (upper panel, middle). The average value of interferon-gamma halved 111 days after second vaccination (95 % prediction interval: 80 to 190 days), and the antibody neutralization capacity after 199 days (95 % prediction interval: 189 to more than 232 days).

A division of the study participants into those with and without co-morbidities showed a similar picture as in the analysis of the antibodies. Co-morbidities of any kind led to reduced T-cell reactivity. It is therefore plausible to assume that the duration of the protective effect against SARS-CoV-2 infection is also limited. Our data show and confirm the data of others that the immune response after two vaccinations varies greatly from individual to individual, but clearly diminishes within the observation period of up to 232 days.

### Correlate of protection

For many technical and medical reasons, there is currently no immunological parameter that allows a reliable statement about the protection status against COVID-19 disease for the individual. Although numerous studies suggest a strong correlation between neutralizing antibody levels and protection (11), many of the regulatory agencies state that antibody tests should not be used to evaluate a person’s level of immunity or protection from COVID-19. This is highly unsatisfactory. At present, a person living in Germany, who has recovered from COVID 19 more than three months ago, has many restrictions in everyday life, irresprective of documented high antibody levels and high IGRA levels. We have therefore analysed the data collected here to estimate what the course of protection would be depending on the time passed after the second vaccination. For this purpose, we defined, according to available evidence (see in the discussion), that a SARS-CoV-2 S1 reactive T cell value above 200 mIU/ml and a neutralisation value of more than 75 % indicates protection against a severe infection.

Based on these assumptions, it appears that the vast majority of persons (95 %) can be assumed to be protected three weeks after the second vaccination, although the small sample size at that observation time causes quite large uncertainty, as indicated by the wide 95 % prediction interval (**Figure 4**). The proportion of protected individuals decreases continuously over time. Fifty percent of persons are still protected 106 days after second vaccination. Younger individuals under 65 years are protected for a longer time; on average, it takes 164 days until only 50 % are still protected, while the proportion of protected older individuals is always below 50 %. Those without comorbidity are protected to 50 % for 149 days after second vaccination and those with comorbidity for 81 days.

**Figure 4:**
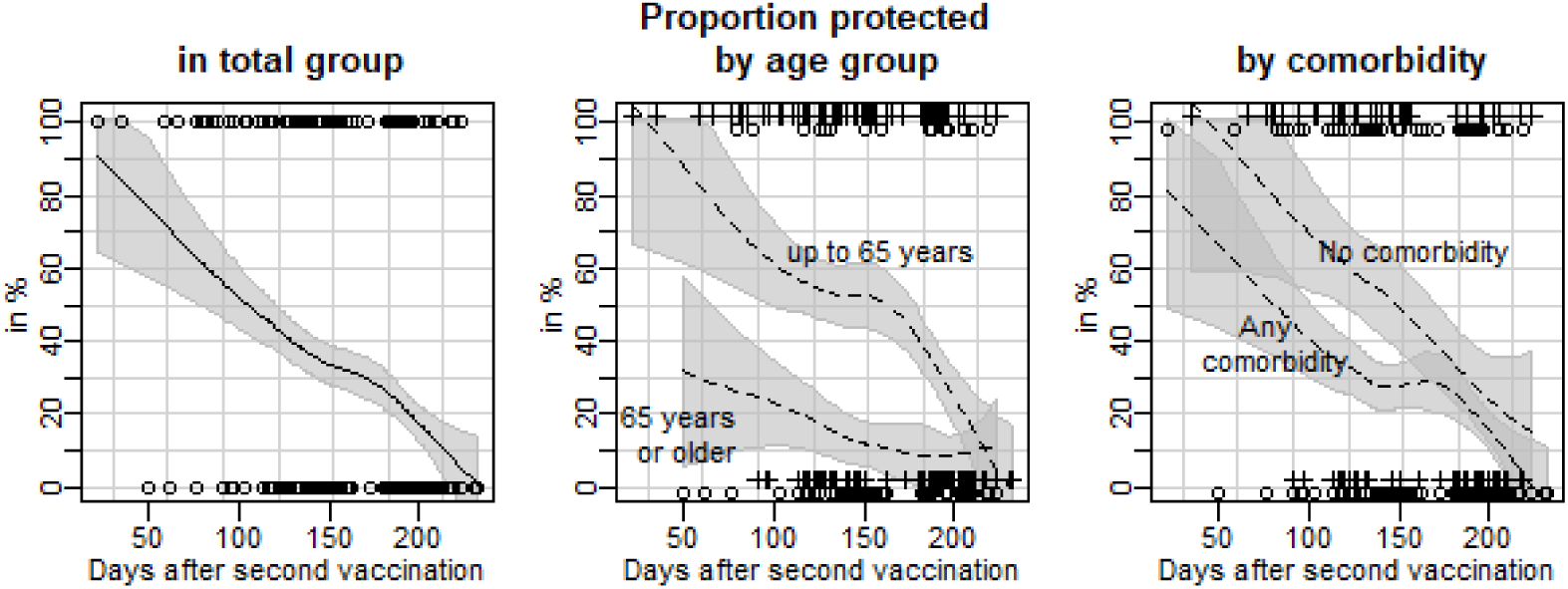
Proportion of persons that are protected against Covid-19 disease over time after second vaccination in the total group, by age group and by comorbidity status. A person is considered to be protected against COVID-19 if the SARS-CoV-2 S1 reactive T cell test is positive, i.e. > 200 mIU/ml, and the neutralisation capacity is > 75 %. Dots and crosses indicate the individual protection status (0 % = not protected, 100 % = protected), line – predicted proportion, grey area – 95 % prediction band

## Discussion

Many observational studies in which the course of the vaccine efficacy of the COVID-19 vaccines is analaysed show, that over a period of 4-6 months after completion of the basic immunisation there is only a slight decline in efficacy against severe COVID-19 disease (hospitalisation). The decline in efficacy against symptomatic infections of any severity, on the other hand, is more pronounced in most studies and amounts to between 10 and 50 % (depending on the vaccine and age group) (12). In consequence, a third vaccination, usually 3 months after the second vaccination, is recommended in most countries to booster the immune system (13).

In this study, we compared the humoral and cellular immune response after two vaccinations of residents of nursing homes over 65 years of age with that of equally vaccinated under-65-year-old employees in these facilities. At the time of the study, the delta variant prevailed. During the study period, no symptomatic breakthrough infection was recorded.

We were able to show that, on average, the initial immune responses in the younger ones were about 30 % higher than in the older ones. Over time, all parameters dropped continuously in all groups within the maximum observation period of 232 days. The existence of any co-morbidities such as coronary heart disease or diabetes mellitus reduced the initial immune responses, regardless of age. These data support and extend the findings from Delbrück et al. (7) and clearly demonstrate the need for a third vaccination. Interestingly, in contrast to an almost linear decline in antibody levels, we observed that the interferon-gamma response remained at a constant level between about day 120 and 180, only to decline further thereafter (**Figure 3**). This might reflect a twofold reaction from the T cell compartment. In the first wave after vaccination, primary T cells with limited longevity are stimulated to produce interferon-gamma, followed by a second wave of long-lived T memory cells that compensate (between 120 and 180 days) for the further drop in interferon-gamma levels (14).

Our data thus show that measurable immune parameters decline within months, accordingly resulting in increasing risks for breakthrough infections in health care workers and in the general population (15). Little is known about what the measurable immunological values mean for the protection of the individual.Therefore, we would like to make a suggestion based on published results and our data.

### Proposal for an immune correlate of protection

Although most of the currently accepted correlates of protection are based on antibody measurements, there is currently no validated immune correlate of protection from SARS-CoV-2 infection, albeit it is urgently needed (16, 17). However, an association between anti-S1 RBD IgG and neutralization antibody levels after immunization with BNT162b2 has been reported (11, 18-20). Correlates may differ depending on the endpoint used, such as protection from infection, from disease, from severe disease or from mortality. At present, the messaging from regulatory agencies states that antibody tests should not be used to evaluate a person’s level of immunity or protection from COVID-19 (21). Having an established correlate of protection would allow healthcare providers to manage, for example, the vaccination of immunocompromised individuals, such as by recommending personalized booster vaccinations or recommending non-pharmaceutical interventions for protection if no immune response is detected.

It would also allow healthcare and governing bodies to efficiently determine what percentage of the population is protected. Although seroprevalence is currently used as a crude measure of community immunity, having a correlate of protection would allow more precise estimations that could then trigger interventions such as vaccination campaigns if the percentage of immune individuals is deemed to be too low.

Finally, there are many persons, who are recovered from COVID-19 and have high antibody levels for a long time. Under the rules currently in force, they are subject to many restrictions, as they do not receive a protection card that allows them to participate in public or even to travel. To avoid the restrictions, they are forced to get vaccinated, which causes a big societal debate. Chau et al. (22) report a median of 91.1 % and an interquartile range of 77.3 % to 94.2 % for neutralizing antibody levels in vaccinated individuals who remained uninfected. (For simplicity, the 77.3 % can be rounded to 75 %.). Feng et al. (11) found that an IgG antibody level of at least 264 BAU/ml is associated with an 80 % vaccine efficacy against primary symptomatic COVID-19. For T-cell parameters, we so far found no studies making a link to protection. With our data (**Figure 4**) and after considering these and other studies, we would like to suggest which people should be considered protected based on immunological values.

We propose to consider persons protected who have a neutralizing capacity of 75 % (which corresponds to approx. 200 BAU/ml) and an interferon-gamma response above 200 mIU/ml. This adoption assumes a normal und functional immune system. A one-time measurement can necessarily only be a snapshot. To estimate the continuity of the protection status, the examinations must be repeated regularly, approximately at intervals of two to three months. For the proportion of protected persons, it is less important to use a neutralization capacity > 75% or the SARS-CoV-2 S1 protein IgG antibody level is > 200 BAU/ml (**Figure 2 in the supplement**). In practice, analysis of the T-cell response may not be necessary (**Figure 3 in the supplement**). Conversely, we were able to show that a higher threshold affects the estimated proportion of people protected (**Figure 4 in the supplement**).

We are very aware that immunological tests are subject to a number of limitations. Although manifold evidence suggests that there is a correlation between neutralising activity in plasma and protection from symptomatic infection at the population level, the titres of neutralising antibodies decrease over time after infection or vaccination; the kinetics of the decrease vary from person to person. Even normalisation to the WHO standard may not fully compensate for the inter-assay variability of pseudovirus-based neutralisation assays. High speed development of variants (of concern) with presumably altered surface properties makes prognosis of protection somewhat difficult, e.g. shift from delta to omicron basically led to a shift from protection against infection” to (limited) protection against severe sequelae/ severe disease”. Consequently this protection correlate might (as an absolute value) only hold true for conditions present during the study period. Furthermore, exposure to high viral loads would require higher protective titres than exposure to low viral loads (e.g. when masks are worn).

### Conclusions

Our study makes statements from a time when the Delta variant was predominant. The conclusions do not necessarily apply to other variants such as Omikron. For residents, the mean time between the second vaccination and blood collection was 22 days longer than for HCW (195 days and 173 days, respectively). This could influence the results in such a way that the decline of SARS-CoV-2 S1 reactive T cells and of the neutralisation capacity over time may be overestimated. On the other hand, this bias may be counteracted by our conservative approach to deal with the summary categories for laboratory results above certain thresholds. We substituted such observations with continuous values very close to the respective threshold. It can be assumed that the truncated values are in truth higher and that consequently reductions over time would be more pronounced (sensitivity analyses not shown). Despite the limitations of our study, it is time to shift the consideration from the population level to the individual. Therefore, this proposal is intended as a stimulus for discussion and needs to be verified by large clinical trials. However, we think that the proposed values are suitable for everyday use.

## Data Availability

All data produced in the present study are available upon reasonable request to the authors.

## Abbreviations

COVID-19: coronavirus disease 19
ELISA: enzyme-linked immunosorbent assay
IgG/A: immunoglobulin G/A
rtPCR: real-time polymerase chain reaction
SARS-CoV-2: Severe acute respiratory syndrome coronavirus 2

## Author contributions

JS, NE and HB had full access to all the data in the study and take responsibility for the integrity of the data and the accuracy of data analysis. SJ, KW, SS, CE and DB were responsible for blood collections and appropriate pre-analytic procedures. DB provided the data of the suitable study participants. BT was responsible for laboratory testing. WS and AM developed the study design and supervised the study. All authors contributed to the drafting of the manuscript and agreed with the final version.

