## Supplemental figures for "Immune responses after twofold SARS-CoV-2 immunisation in elderly residents and Health Care Workers in nursing homes and homes with assisted living support - Proposal for a correlate of protection"

**Supplemental Figure 1:** Pairwise bivariate scatterplots and Spearman correlation coefficients for Anti-SARS-CoV-2 S1-Protein IgG Antibodies,
neutralisation capacity and Interferon-gamma

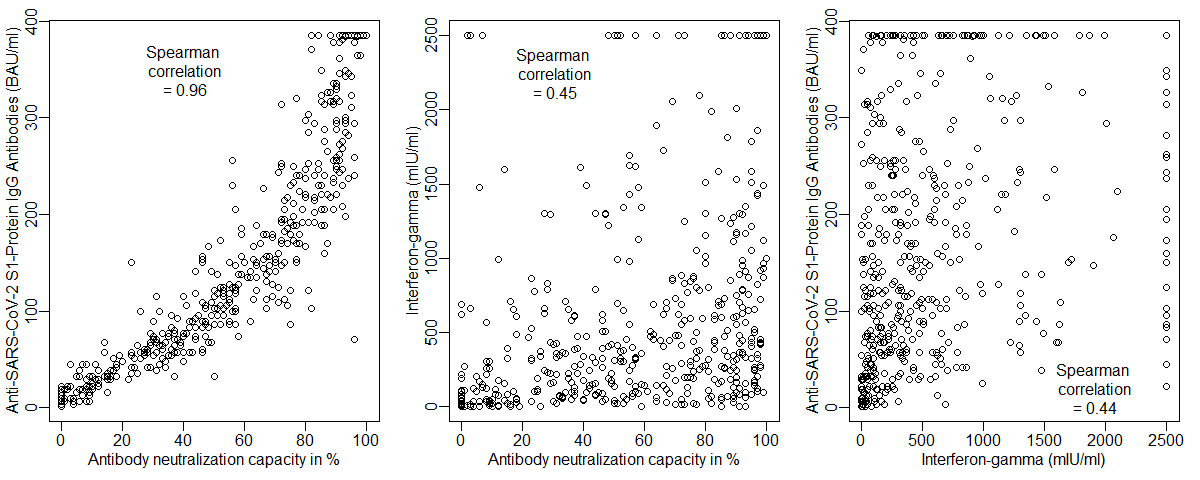

**Supplemental Figure 2:** Proportion of persons that are protected against Covid-19 disease over time after second vaccination in the total group, by age group and by comorbidity status.

A person is considered to be protected against COVID-19 if the SARS-CoV-2 S1 reactive T cell test is positive, i.e. > 200 mIU/ml, and the Anti-**SARS-CoV-2 S1-Protein IgG Antibody level is > 200 BAU/ml** (which corresponds to a neutralisation capacity of > 75 %).

Dots and crosses indicate the individual protection status (0 % = not protected, 100 % = protected), line – predicted proportion, grey area – 95 % prediction band

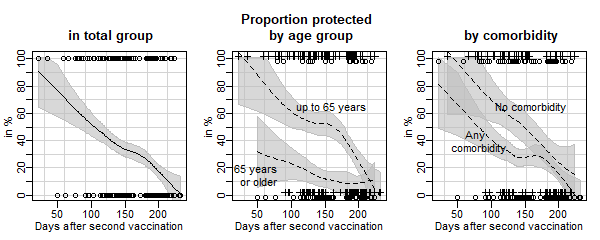

**Supplemental Figure 3:** Proportion of persons that are protected against Covid-19 disease over time after second vaccination in the total group, by age group and by comorbidity status.

A person is considered to be protected against COVID-19 if only the Anti-**SARS-CoV-2 S1-Protein IgG Antibody level is > 200 BAU/ml** (which corresponds to a neutralisation capacity of > 75 %).

Dots and crosses indicate the individual protection status (0 % = not protected, 100 % = protected), line – predicted proportion, grey area – 95 % prediction band

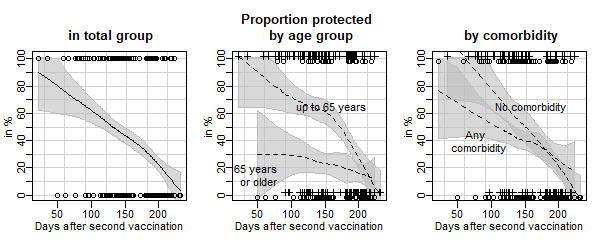

**Supplemental Figure 4:** Proportion of persons that are protected against Covid-19 disease over time after second vaccination in the total group, by age group and by comorbidity status.

A person is considered to be protected against COVID-19 if the Anti-**SARS-CoV-2 S1-Protein IgG Antibody level is > 264 BAU/ml** (threshold from Feng et al. (11)).

Dots and crosses indicate the individual protection status (0 % = not protected, 100 % = protected), line – predicted proportion, grey area – 95 % prediction band

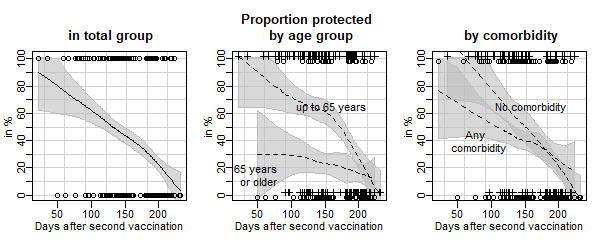
